# Systematic Integration of genomics with transcriptomics for the Study of Coronary Artery Disease and Subclinical Atherosclerosis

**DOI:** 10.64898/2026.07.31.26357396

**Authors:** Chaojie Yang, Francois Aguet, Gaelle Auguste, Kristin Ardlie, Robert Gerszten, Wendy S. Post, Heather E. Wheeler, Kent D. Taylor, Silva Kasela, Tuuli Lappalainen, Hae Kyung Im, Peter Durda, Craig Johnson, Xiuqing Guo, Yongmei Liu, Joseph Polak, David Herrington, Clary Clish, David Van Den Berg, Russell P. Tracy, Elaine Cornell, Tom Blackwell, George Papanicolaou, Jose D. Vargas, Stefan Bekiranov, Coleen A. McNamara, Clint L. Miller, Jerome I. Rotter, Stephen S. Rich, Ani Manichaikul

**Author notes:** **Correspondence:** Ani Manichaikul, PhD, Associate Professor of Genome Sciences, Department of Genome Sciences, University of Virginia School of Medicine, West Complex Room 6115, Charlottesville, VA 22903.

## Abstract

**Introduction:** Coronary artery disease (CAD) is a leading cause of death and disability worldwide. Although genome-wide association studies (GWAS) have identified over 300 loci associated with CAD risk, the molecular mechanisms linking these variants to disease and subclinical atherosclerosis are not fully understood.

**Methods:** We performed integration of multi-ancestry CAD GWAS with transcriptomic data from the Multi-Ethnic Study of Atherosclerosis (MESA) obtained through the Trans-Omics for Precision Medicine (TOPMed) program. For integration, we applied Bayesian colocalization analysis with and without statistical fine-mapping to identify genes whose expression levels colocalize with CAD-associated loci. We further applied causal weighted gene co-expression network analysis (cWGCNA) to identify gene co-expression modules and key driver genes associated with subclinical atherosclerosis traits in MESA.

**Results:** We identified 108 genes showing evidence of colocalization with CAD loci, including 24 shared between the two colocalization approaches and 48 novel genes not previously reported in CAD GWAS. Follow-up replication and validation analyses prioritized 5 novel (*CCDC30, ZEB1-AS1, ZPR1, PLEKHJ1 and AC018816.3*) and 8 previously reported genes (*DHDDS, DDX59, LNPEP, DAGLA, ZKSCAN1, LIPA, OPRL1 and EIF2B2*) with putative roles in both CAD and subclinical atherosclerosis. cWGCNA identified five gene modules significantly associated with subclinical atherosclerosis in MESA. Additionally, three key driver genes (*ATG9B, PRAM1* and *ZBTB46*) identified by cWGCNA were also identified as CAD-colocalized genes.

**Discussion:** Our integrative analysis highlights key genetic drivers and regulatory networks underlying CAD and subclinical atherosclerosis. These findings underscore the value of incorporating statistical fine-mapping in colocalization studies and demonstrate the utility of combining colocalization with co-expression network analysis to prioritize functional genes and pathways.

## Introduction

Coronary artery disease (CAD) is a common complex disease with both genetic and environmental determinants and is a leading cause of death and disability worldwide.^1,2^ The primary cause of CAD is atherosclerosis, characterized by the progressive buildup of plaques composed of lipids, calcium, fibrin, and inflammatory cells within the arterial wall.^3–7^ Notably, quantification of coronary artery calcification (CAC) via computer tomography (CT) calcium scoring is a widely recognized marker of subclinical atherosclerosis. Similarly, the presence of carotid plaque and increased carotid intima-media thickness (IMT) are significantly associated with CAD, and together, these traits reflect early vascular changes preceding clinical cardiovascular events.^8–12^ Risk factors contributing to CAD susceptibility can be categorized into two primary categories: non-modifiable and modifiable determinants. Non-modifiable risk factors include male sex, a familial history of premature heart disease and advanced age. In contrast, modifiable risk factors are amendable to intervention through lifestyle adjustments and medical management. These factors include cigarette smoking, dyslipidemia, elevated blood pressure, diabetes, excess body weight or obesity, and an unhealthy diet.^13–15^ Understanding how these risk factors intersect with genetic susceptibility is key to improving prevention and treatment strategies.

Genetic studies have substantially advanced our understanding of the genetic basis of CAD.^16^ GWAS have identified over 300 independent loci associated with CAD, with most findings initially derived from populations of European ancestry.^17,18^ More recently, a trans-ethnic GWAS by Tcheandjieu et al. using data from the Million Veteran Program (MVP), UK Biobank, CARDIoGRAMplusC4D, and Biobank Japan, identified 95 novel CAD-associated loci across diverse populations.^19^ Despite these successes, the downstream molecular consequences of CAD-associated variants remain largely uncharted in human cohorts, particularly in relation to subclinical atherosclerosis.

To bridge this knowledge gap, it is imperative to perform integrative analyses by leveraging multi-omics data including transcriptomics, proteomics, metabolomics, and epigenomics, to identify the molecular targets that causally linked to CAD and provide critical guidance in enhancing the accuracy of disease diagnosis and prognosis. ^20,21^ Resources from the Trans-Omics for Precision Medicine (TOPMed) Multi-Ethnic Study of Atherosclerosis (MESA) offer multiple molecular ‘omics datasets, including RNA-seq from peripheral blood mononuclear cells (PBMCs), CD4+ T cells, and monocytes, genome-wide DNA methylation from whole blood, and plasma proteomics measured via SOMAscan. Additionally, MESA participants have deep phenotyping available, including imaging-based measures of subclinical atherosclerosis, providing a unique resource for examination of disease relevance for identified molecular targets.^22,23^ Accordingly, focused investigation of molecular targets implicated by molecular measures for circulating samples from the MESA participants can provide a valuable complement to ongoing investigations that make use of gene expression on coronary artery and other disease proximal tissues.^24,25^

Bayesian colocalization analysis, has proven a rigorous and efficient computational approach for identification of downstream molecular targets underlying GWAS loci. Colocalization analysis quantifies the probability that a single variant is causally linked to both disease and molecular traits, which in turn suggests that the gene regulated by this variant is a candidate gene for the observed trait.^26^

Colocalization analysis has been widely used in various research studies to identify candidate genes associated with complex traits and diseases.^27, 28^ More recently, the incorporation of statistical fine-mapping into colocalization analysis represents a more flexible approach for the identification of candidate genes underlying the GWAS loci, allowing for multiple independent causal variants within a single locus.^29^

In this study, we aimed to prioritize candidate genes and pathways involved in CAD and subclinical atherosclerosis by integrating multi-ancestry GWAS^19^ with transcriptomic data from TOPMed MESA. We performed Bayesian colocalization analysis - with and without statistical fine-mapping - to identify CAD colocalized genes using eQTLs from multiple immune cell types. We further performed multiple follow-up analyses to investigate the relationship of CAD-colocalized genes with subclinical atherosclerosis and prioritize a list of candidate causal genes of CAD and subclinical atherosclerosis. We additionally conducted causal weighted gene co-expression network analysis (cWGCNA) to identify transcriptional modules associated with subclinical atherosclerosis traits (CAC, IMT, and carotid plaque) in MESA and further prioritize the key driver within subclinical atherosclerosis related modules **(Figure 1).** Overall, our study combines integrative genomic and transcriptomic approaches with network-based analysis to prioritize candidate causal genes contributing to CAD and subclinical atherosclerosis and provide mechanistic insights into disease pathogenesis.

**Figure 1.**
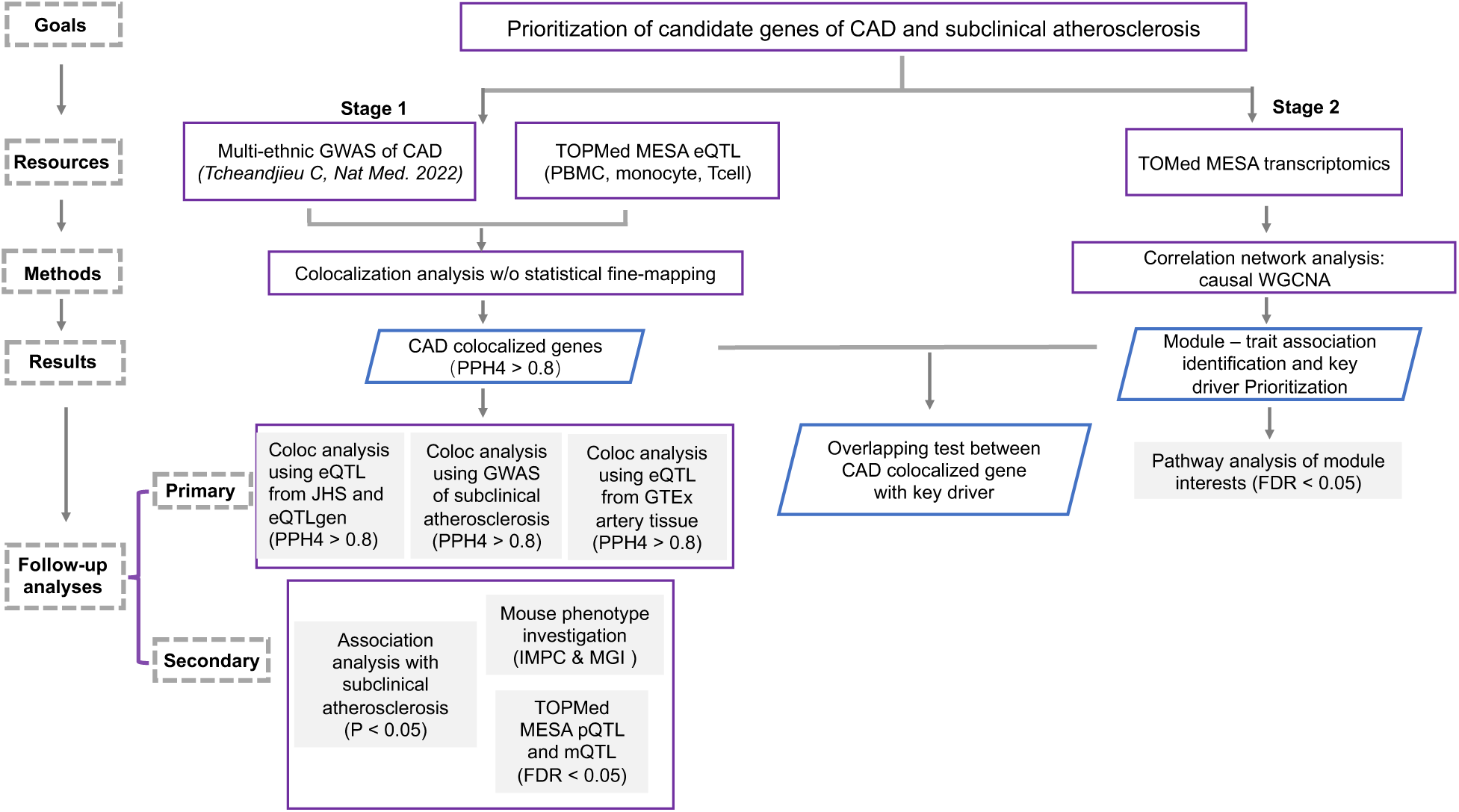
Study design. Colocalization analysis w/o statistical fine-mapping were performed to identify the colocalized gene of CAD by using GWAS of CAD and TOPMed MESA eQTL. Primary follow-up validation analyses include (a) exploration of the colocalized genes in GWAS of subclinical atherosclerosis, (b) investigation of colocalized genes in Artery tissue from GTEx and (c) exanimation of the colocalized genes in PBMC from JHS and whole blood from eQTLgen and secondary follow-up validation analyses include (a) examination of the association of colocalized genes with subclinical atherosclerosis traits in MESA, (b) examination of causal CAD variants for evidence of pQTL and mQTL associations for the corresponding colocalized genes in TOPMed MESA, (c) study of the colocalized genes in mouse genome. Causal weighted gene co-expression network analysis (cWGCNA) was conducted to identify gene expression modules and investigate their association with subclinical atherosclerosis traits in MESA and examined the modules identified for enrichment of the colocalized genes identified by Bayesian colocalization analysis.

## Methods

### Multi-Ethnic Study of Atherosclerosis (MESA) and MESA Multi-omics: Study design

MESA is a longitudinal cohort study of subclinical cardiovascular disease and risk factors that predict progression to clinically overt cardiovascular disease or progression of subclinical disease. Between 2000 and 2002, MESA recruited 6,814 men and women 45 to 84 years of age, free of clinical cardiovascular disease, from Forsyth County, North Carolina; New York City, New York; Baltimore, Maryland; St. Paul, Minnesota; Chicago, Illinois; and Los Angeles, California. Participants at baseline were 38% White, 28% African American, 22% Hispanic and 12% Chinese ancestry.^30^

### Primary analysis for identification of CAD colocalized genes

#### TOPMed MESA transcriptomics and eQTL

As part of the TOPMed MESA Multi-Omics project, participants were selected for transcriptome profiling based on the following criteria: (1) restrict to those already included in the TOPMed Whole Genome Sequencing effort ^31^, (2) preserve the race/ethnic distribution of participants in the parent MESA cohort, (3) maximize the amount of overlapping ‘omics data (with the other ‘omics included in the TOPMed MESA Multi-Omics pilot requiring availability of plasma samples for proteomics/metabolomics and whole blood for genome-wide methylation). MESA transcriptomics and cis-eQTL were obtained through TOPMed Freeze 1RNA. Freeze 1RNA TOPMed cis-eQTL results were generated in a collaboration between the TOPMed Informatics Research Center, TOPMed Multi-Omics working group, and the TOPMed parent studies contributing RNA-seq and distributed to TOPMed investigators. The cis-eQTL mapping was performed in MESA exam5 using tensorQTL^32^ for PBMCs (n = 1256), T cells (n = 368) and monocytes (n = 352), and for each gene, genetic variants within 1Mb of the gene TSS were tested. The covariates for cis-eQTL mapping included sex, 15 genotype PCs and 30 gene expression PCs.

Samples for the cis-eQTL scan were selected via the following procedure: (1) exclude samples that did not pass the PC-based outlier filter, (2). exclude samples without a known WGS match, (3). exclude samples where the subject (WGS match) is not in the unrelated subject set, and (4) exclude samples with unclear sex based on gene expression. In the MESA transcriptomic dataset used in this study, samples were processed within a single batch.

#### GWAS of coronary artery disease (CAD)

We leveraged a recently published publicly available large-scale multi-ancestry of GWAS of CAD comprising of 243,392 cases and 849,686 controls.^41^ This study used METAL to conduct a fixed-effect inverse variance-weighted meta-analysis for the clinical CAD phenotype, including Million Veteran Program (MVP) European participants MVP Black participants, MVP Hispanic participants and Biobank Japan, CARDIoGRAMplusC4D 1000G study, the UK Biobank CAD study and Biobank Japan.

#### Statistical fine-mapping for identification of causal variants of GWAS and eQTL

We carried out statistical fine-mapping using Sum of Single Effect model (SuSiE^33,34^) to identify the putative causal variants and estimate the number of independent signal, leveraging the summary statistic results including effect size, standard error and minor allele frequencies in GWAS of CAD and MESA eQTL, with L = 10 (SuSiE default). Each credible set was constructed to have a high probability of containing a variant with a non-zero effect, while being as small as possible. Specifically, we defined a window size of ±0.5 Mb of the transcriptional start sites of each gene in the MESA eQTL and apply SuSIE to identify the credible set of putative causal variants within this window for both the GWAS of CAD and MESA eQTL. Subsequently, the follow-up colocalization analysis was conducted to systematically evaluate all potential pairs of credible sets between the GWAS and eQTL results.

#### Bayesian colocalization analysis for identification of colocalized genes underlying CAD GWAS loci

Bayesian colocalization analysis^26,29^ was used to identify the downstream genes of CAD leveraging MESA eQTL and GWAS of CAD by using R/coloc package. Colocalization analysis quantifies the probability that a single variant is causally linked to both CAD and molecular traits, with the following hypotheses: H0. neither GWAS nor eQTL has a genetic association in the region; H1. only GWAS has a genetic association in the region; H2. only eQTL has a genetic association in the region; H3. both GWAS and eQTL are associated, but with different causal variants; H4. both GWAS and eQTL are associated and share a single causal variant. A posterior colocalization probability of hypothesis 4 (PP.H4) > 0.80 was used as the threshold of colocalization. A distinguishing feature of this approach is the minimal input data requirement, which includes p-values of the trait-associated SNPs and their minor allele frequencies (MAFs), or effect size of SNPs and the corresponding standard error. The credible sets of causal variants identified by SuSIE for both CAD GWAS and MESA eQTL within the same genomic region were systematically evaluated through the colocalization analysis to identify CAD colocalized genes. Both colocalization analysis with/without statistical fine-mapping were applied on MESA PBMC eQTL and GWAS of CAD colocalization analysis. The primary colocalization analysis in this study was performed by integrating CAD GWAS summary statistics with eQTL data derived from MESA PBMCs. Additional colocalization analyses using eQTLs from CD4+ T cells and monocytes were conducted as sensitivity analyses to evaluate the cell type specificity of the identified regulatory signals and to assess whether prioritized genes were shared across immune cell populations or enriched within specific cell types.

### Identification of novel versus previously reported CAD colocalized gene

To distinguish novel versus previously reported CAD colocalized genes, we utilized results from gene-based analyses in a previously published CAD GWAS^19^. The gene-based analyses include DEPICT, MAGMA, RSS-E and MetaXcan. For each CAD colocalized genes identified in the present colocalization analysis, those reported in any of the previous gene-based analyses were considered known, while the remaining genes were considered novel in the current study.

### Follow-up validation approaches

Focusing on the CAD colocalized genes from primary colocalization analysis using GWAS of CAD and MESA PBMC eQTL, primary and secondary follow-up validation analyses were performed to further prioritize the 108 candidate genes identified by colocalization between GWAS of CAD and MESA PBMC eQTL.

Primary follow-up validation analyses for the 108 genes colocalized between GWAS of CAD and MESA PBMC eQTL included additional colocalization analyses for to examine evidence of shared causal variants for (1) GWAS of subclinical atherosclerosis with MESA PBMC eQTL to examine generalizability to the identified genes to atherosclerosis, (2) GWAS of CAD with GTEx eQTL from artery tissue to examine overlap of the identified signals with disease proximal tissue, and (3) GWAS of CAD with PBMC eQTL from the Jackson Heart Study (JHS) and whole blood eQTL from eQTLGen to examine reproducibility of the identified colocalization relationships based on independent sources of eQTL from circulating samples.

Secondary follow-up validation of the 108 CAD colocalized genes included (1) examination of association of measured gene expression with subclinical atherosclerosis traits in MESA, (2) investigation of fine-mapped CAD variants for evidence as pQTL for the proteins corresponding to the CAD colocalized genes and mQTL for methylation sites within regions (±0.5 Mb of the transcriptional start sites) for the CAD colocalized genes, and (3) *in silico* examination of the effects of the identified genes on disease relevant phenotypes in mouse.

### Primary follow-up validation analysis

#### Colocalization analysis for GWAS of subclinical atherosclerosis with MESA PBMC eQTL

To investigate the relationship between CAD-colocalized genes with subclinical atherosclerosis, standard colocalization analysis focusing on the CAD colocalized genes from primary analysis was performed to identify the downstream genes of subclinical atherosclerosis leveraging MESA eQTL and GWAS of subclinical atherosclerosis (CAC^35^ and IMT^36^) by using R/coloc package (coloc.abf). For GWAS of CAC, we leveraged a published publicly available meta-analysis GWAS of CAC comprising 9,961 participants from 5 independent community-based cohorts (Age, Gene/Environment Susceptibility–Reykjavik Study [AGES-Reykjavik], the Framingham Heart Study [FHS], the Rotterdam Study I [RS I], and the Rotterdam Study II [RS II], Genetic Epidemiology Network of Arteriopathy Study [GENOA]).^35^ For GWAS of IMT, we leveraged a published publicly available meta-analyses of GWAS of IMT in 71,128 individuals of European ancestry from 31 studies for IMT.^36^

#### Investigation of CAD colocalized genes in artery tissue from GTEx

Even though, our primary analysis leveraged MESA whole blood eQTL, arterial tissue is more directly related to the CAD phenotype. To examine the effect of CAD-colocalized genes in arterial tissue, we leveraged GTEx eQTL^37^ to perform standard colocalization analyzes using R/coloc package (coloc.abf). The GTEx eQTL used in colocalization analysis included coronary artery tissue (n = 213), aortic tissue (n = 387) and tibial artery tissue (n = 584). GTEx eQTL can be found and downloaded at https://www.gtexportal.org/home/downloads/adult-gtex.

#### Exanimation of the colocalized genes in PBMC from Jackson Heart Study (JHS) and whole blood from eQTLGen

To replicate our CAD colocalized genes using eQTL from other circulating samples (PBMCs or whole blood), standard colocalization analysis (coloc.abf) was performed using eQTL resources from JHS and eQTLGen. JHS-eQTL was generated from 1,012 African Americans from the Jackson Heart Study by leveraging their PBMC RNA-sequencing data (∼50 million reads per sample) and whole genome sequencing (WGS) data from the NHLBI Trans-Omics for Precision Medicine (TOPMed) program.^31^ JHS-eQTL contains a total of 4,524,238 variant-gene pairs significant at FDR 10% including 16,670 unique genes. The eQTLGen cis-eQTL dataset was generated from whole-blood gene expression data and genotype data from 31,684 individuals across 37 cohorts and identified significant cis-eQTLs for 16,987 genes (88% of tested genes) at FDR < 0.05.^38^

### Secondary follow-up validation analysis

#### Examination of association of measured gene expression with subclinical atherosclerosis traits in MESA

CAD-colocalized genes identified from primary analysis were carried forward to examine the association with subclinical atherosclerosis traits (CAC, IMT and carotid plaque) in MESA exam 5 using linear regression model and the covariates included age, sex and study sites.

#### Examination of causal CAD variants for evidence of pQTL and mQTL associations for the corresponding colocalized genes

shared causal variants identified from colocalization analysis with statistical fine-mapping were further investigated to assess their potential impact as MESA pQTL or MESA mQTL for the proteins and CpG sites corresponding to CAD-colocalized genes.

#### Investigation of CAD colocalized genes in mouse genome

To investigate the biological function of CAD-colocalized genes in mouse genome, we utilized the data from the International Mouse Phenotyping Consortium (IMPC)^39^ and Mouse Genome Informatics (MGI)^40^ to determine whether these CAD-colocalized genes are associated with heart/cardiovascular phenotypes Detailed methods for the secondary follow-up validation approaches are included in the Supplementary Methods text.

### Single-nucleus ATAC-seq (snATAC) analysis

We leveraged coronary artery snATAC-seq data^41^ to (1) query the potential causal genes of CAD from colocalization analysis and WGCNA to check whether they were identified as cell type specific marker gene and (2) investigate whether any of potential causal variants identified from statistical fine-mapping overlap peak-to-gene links. The variant positions were checked for intersection with snATAC peaks that were unique to a specific coronary-artery cell-type using *bedtool* intersect (version 2.30.0,) using the defaults parameters. The correlations between co-accessible variants and gene expression were assessed using the *ArchR* peak2genes-based predictions.^42^ Both analyses were performed after increasing the genomic interval to a 100-bp window centered on the variants. The genes from the colocalization analysis and the hub genes from WGCNA were then individually checked against the list of marker genes across all coronary artery cell types as defined in Turner et al., in R (version 4.1.1). Cell-type specific marker genes in coronary artery snATAC data (genes with significantly higher chromatin accessibility in a cluster than in other clusters) were identified using Wilcoxon rank-sum test and the genes with (Benjamini-Hochberg) adjusted p-value <= 0.01 and fold change >=2 were selected.

### Causal weighted gene co-expression network analysis (cWGCNA)

Causal weighted gene co-expression network analysis (cWGCNA) is a methodological extension of the traditional WGCNA framework that aims to infer potential causal relationships among gene co-expression modules, individual gene expression features, and phenotypic traits while accounting for confounding factors. By integrating causal mediation analysis into the co-expression network framework, cWGCNA enables the prioritization of modules and genes that are more likely to be causally associated with phenotypic variation and this approach provides a principled framework for improving the causal interpretability of co-expression networks in studies of complex diseases. Our study applied cWGCNA on MESA exam 5 PBMC transcriptomic data including 1,256 unique individuals and 24,410 genes to identify subclinical atherosclerosis traits (CAC, IMT and carotid plaque) related modules. We performed analysis of variance (ANOVA) across available covariates, including study site, race, sex, age and PC, to identify potential confounding factors. A weighted gene co-expression network was then constructed using the WGCNA framework based on pairwise correlations between gene expression profiles, and an appropriate soft-thresholding power was selected to approximate scale-free network topology.

Genes were hierarchically clustered according to topological overlap, and co-expression modules were identified using dynamic tree cutting. Module eigengenes, defined as the first principal component of each module, were subsequently calculated to represent overall module-level expression patterns. Associations between module eigengenes and subclinical atherosclerosis phenotypes in MESA exam5 were evaluated using limma while adjusting for identified confounding covariates. Modules significantly associated with clinical phenotypes after multiple testing correction were prioritized for downstream analyses, and gene-level testing within significant modules was performed to identify highly connected and phenotype-associated candidate driver genes.^43^

### Pathway enrichment analysis for identification of biological pathways

We applied Gene Set Enrichment Analysis (GSEA) to investigate the biological pathways for the genes within the module of interest identified by our cWGCNA using the Molecular Signature Database (MSigDB).^44,45^ MSigDB includes multiple categories, for example, hallmark gene sets (H), curated gene sets (C2), regulatory target gene sets (C3), computational gene sets (C4), ontology gene sets (C5), oncogenic signature gene sets (C6), immunologic signature gene (C7), cell type signature gene sets (C8).

## Results

### Identification of colocalized genes underlying GWAS of CAD

We performed integration of summary statistics from a GWAS of CAD^19^ with PBMC eQTL data from the TOPMed MESA cohort by applying Bayesian colocalization analysis. A total of 108 colocalized genes were identified across the colocalization analyses conducted with and without statistical fine-mapping. Specifically, 86 genes were identified without fine-mapping and 46 with fine-mapping, with 24 genes overlapping between the two approaches. **(Figure 2a and Table S1)** Among these colocalized genes, 60 genes had also been previously reported in gene-based analyses of the same CAD GWAS dataset^19^ **(Figure 2a)**, supporting the robustness of our findings. For instance, *PLEKHJ1* is a gene identified by colocalization with CAD GWAS^19^ through analyses both with and without fine-mapping. Fine-mapping successfully identified 3 potential causal variants associated with CAD and 27 potential causal variants linked to *PLEKHJ1* gene expression. Two of these variants are overlapping and exhibit robust associations with both CAD and the expression levels of *PLEKHJ1* (**Figure 2b**). This finding suggests a potential mechanistic link between these causal genetic variants, CAD risk, and the regulation of *PLEKHJ1*.

**Figure 2.**
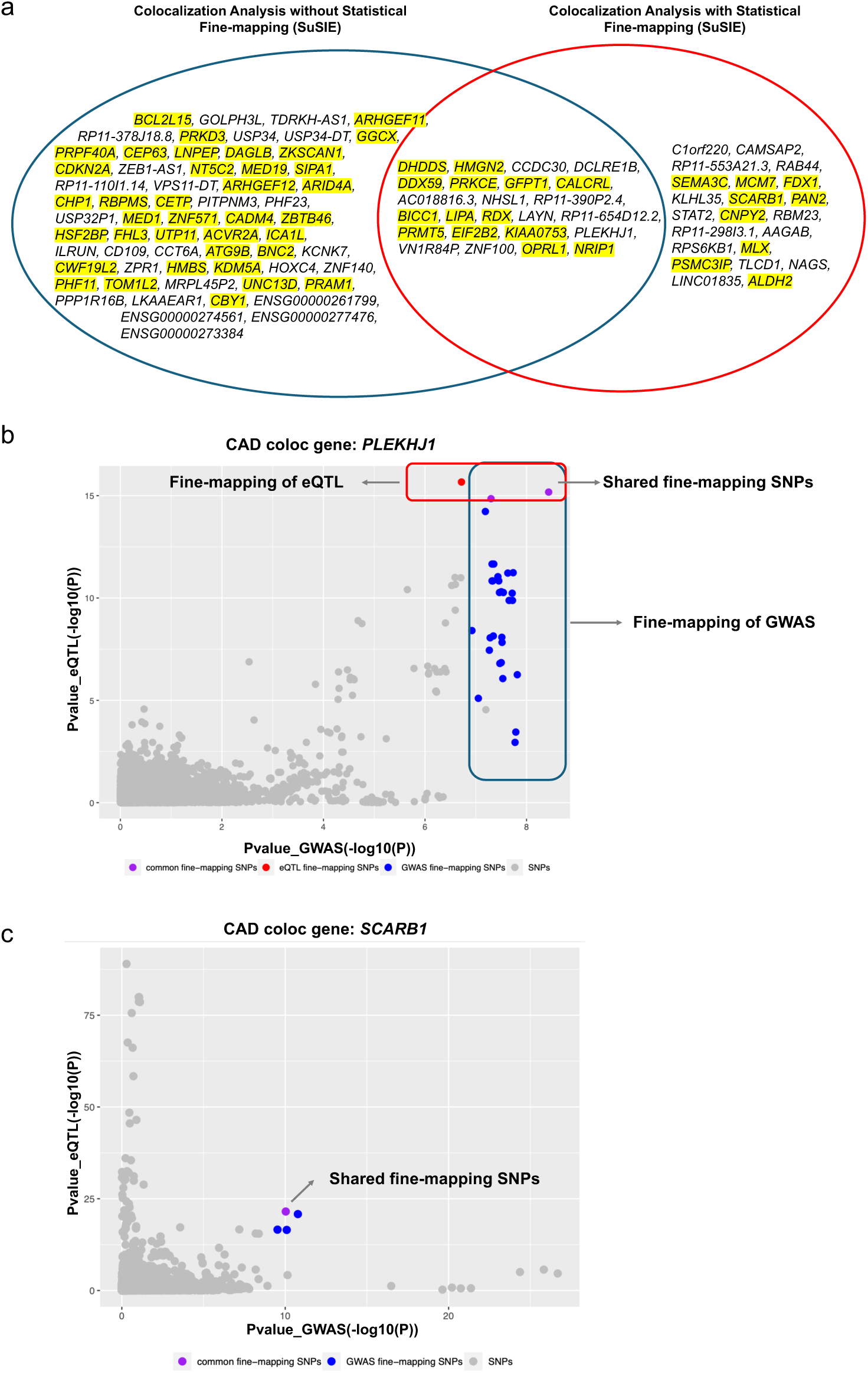
Identification of CAD-colocalized genes by colocalization analysis with and without statistical fine-mapping. (85 genes were identified without fine-mapping and 46 with fine-mapping, with 24 genes overlapping between the two approaches). Highlighted genes shown in **Panel a** had been previously reported in gene-based analyses of the same CAD GWAS dataset. **Panel b and c** show two examples of colocalized genes (*PLEKHJ1* and *SCARB1*). Specifically, *PLEKHJ1* could be identified by both methods, while *SCARB1* could be identified only by colocalization analysis with statistical fine-mapping.

In a different example, *SCARB1* is a colocalized gene that would have remained unidentified without the incorporation of statistical fine-mapping. There were multiple credible sets from both the GWAS of CAD and the eQTL signals within the region of *SCARB1*. After performing colocalization analysis based on each pair of credible sets, one shared causal variant demonstrated strong associations with both CAD risk and expression of *SCARB1*, suggesting the significance of incorporating statistical fine-mapping into the Bayesian colocalization analysis. (**Figure 2c**)

To assess whether CAD colocalized genes identified from primary analysis using PBMC eQTL were shared across immune cell populations or enriched within specific cell types, we extended our colocalization analyses to eQTL data from CD4+ T cells and monocytes in MESA. This revealed 65 and 53 CAD-colocalized genes in CD4+ T cells and monocytes, respectively. In total, 21 genes were consistently colocalized across all three cell types (PBMC, CD4+ T cells, monocytes), while others were cell type–specific (PBMC: 53 unique genes; CD4+ T cells: 22; monocytes: 17). (**Figure S1a and Table S1)** These differences can be attributed to variations in sample sizes of the eQTL data (1,256 participants contributed PBMCs while 368 contributed CD4+ T cells and 352 monocytes**, Figure S1b)**, differences in gene expression profiles across distinct cell types and differences in biological roles among these cell types.

To further evaluate the consistency of our findings with recent CAD GWAS, we compared the lead shared variants identified through our colocalization analyses with results reported by Aragam et al.^46^ Overall, 76 of 108 (70.4%) lead variants remained significantly associated with CAD after Bonferroni correction (*P* < 4.63 × 10^−4^), including the lead variants at the *FHL3* (rs11210866; *P* = 3.46 × 10^−10^) and *DDX59* (rs4387211; *P* = 6.90 × 10^−08^). **(Table S2)**

Additionally, to further interpret the biological relevance of CAD colocalized genes, we examined the direction of effect of the lead shared variants identified through colocalization analyses to evaluate the relationship between genetically regulated gene expression and CAD risk. For example, the lead shared variant at the *ALHD2* locus, rs847892, exhibited a negative effect on CAD risk in both the Tchandjieu et al. CAD GWAS (beta = −0.0337, *P* = 6.14 × 10^−15^) and the Aragam et al. CAD GWAS (beta = − 0.0381, *P* = 6.78 × 10^−13^). In the MESA PBMC eQTL analysis, the same variant was associated with increased *ALDH2* expression (beta = 0.4733, P = 6.28 × 10^−21^), indicating that the allele associated with reduced CAD risk was also associated with increased *ALDH2* expression. **(Table S3)**

### Follow-up validation analyses prioritize the causal genes of CAD and subclinical atherosclerosis

Focusing on the identified CAD-colocalized genes from PBMCs, primary and secondary follow-up validation analyses were performed to prioritize putative causal genes for CAD and subclinical atherosclerosis, yielding the following 13 genes: *DHDDS, CCDC30, DDX59, LNPEP, DAGLA, ZKSCAN1, LIPA, ZEB1-AS1, ZPR1, OPRL1, EIF2B2, PLEKHJ1* and *AC018816.3*. Among these, *CCDC30, ZEB1-AS1, ZPR1, PLEKHJ1 and AC018816.3* represent novel findings not previously identified as candidates in the published CAD GWAS study^19^, while the remaining candidate genes (*DHDDS, DDX59, LNPEP, DAGLA, ZKSCAN1, LIPA, OPRL1 and EIF2B2*) have been previously reported.(**Table 1**) Notably, several prioritized genes also showed evidence of colocalization in CD4+ T cells and monocytes, including *DHDDS*, *CCDC30, DDX59, LIPA, ZEB1-AS1, ZPR1, EIF2B2* and *AC018816.3* in T cells, and *CCDC30*, *DDX59, LNPEP, LIPA OPRL1* and *EIF2B2* in monocytes (**Table S1**), further supporting their potential relevance across multiple immune cell types.

**Table 1.** Prioritization of causal genes for CAD from primary follow-up analyses. These prioritized genes were selected based on the criterion that they showed colocalization with CAD GWAS loci in our primary discovery analysis and passed our specified thresholds in at least two follow-up primary validation analyses. Primary follow-up validation analyses included (a) examination of colocalization with GWAS of subclinical atherosclerosis and (b) investigation of expression artery tissue from GTEx and (c) exanimation of the colocalized genes in PBMC from JHS and whole blood from eQTLgen. Discovery_PPH4: PPH4 generated from the colocalization analysis using GWAS of CAD and MESA PBMC eQTL; Replication_JHS_PPH4: PPH4 generated from the colocalization analysis using GWAS of CAD and JHS PBMC eQTL; Replication_eQTLgen_PPH4: PPH4 generated from the colocalization analysis using GWAS of CAD and eQTLgen PBMC eQTL; CAC_PPH4: PPH4 generated from the colocalization analysis using GWAS of CAC and MESA PBMC eQTL; IMT_PPH4: PPH4 generated from the colocalization analysis using GWAS of IMT and MESA PBMC eQTL; Gtex_Conorary_PPH4: PPH4 generated from the colocalization analysis using GWAS of CAD and GTEx Conorary eQTL; Gtex_ Aorta_PPH4: PPH4 generated from the colocalization analysis using GWAS of CAD and GTEx Aorta eQTL; Gtex_Tibial_PPH4: PPH4 generated from the colocalization analysis using GWAS of CAD and GTEx Tibial eQTL;

| Gene | Discovery_PPH4 | Replication_JHS_PPH4 | Replication_eQTLgen_PPH4 | CAC_PPH4 | IMT_PPH4 | Gtex_Conorary_PPH4 | Gtex_Aorta_PPH4 | Gtex_Tibial_PPH4 | Novel/Reported |
| --- | --- | --- | --- | --- | --- | --- | --- | --- | --- |
| DHDDS | 0.961 | 0.991 | 0.919 | 0.014 | 0.000 | 0.078 | 0.698 | 0.933 | reported |
| CCDC30 | 0.954 | 0.056 | 0.921 | 0.054 | 0.008 | 0.017 | 0.924 | 0.829 | novel |
| DDX59 | 0.991 | 0.992 | 0.886 | 0.015 | 0.027 | 0.984 | 0.983 | 0.001 | reported |
| AC018816.3 | 0.999 | NA | 0.996 | 0.006 | 0.013 | 0.537 | 0.201 | 0.991 | novel |
| LNPEP | 0.929 | 0.954 | 0.894 | 0.188 | 0.017 | 0.054 | 0.907 | 0.922 | reported |
| DAGLB | 0.923 | 0.617 | 0.883 | 0.017 | 0.009 | 0.889 | 0.511 | 0.511 | reported |
| ZKSCAN1 | 0.964 | 0.936 | 0.000 | 0.105 | 0.017 | 0.384 | 0.166 | 0.962 | reported |
| ZEB1-AS1 | 0.965 | 0.943 | 0.464 | 0.013 | 0.007 | 0.618 | 0.867 | 0.881 | novel |
| ZPR1 | 0.851 | 0.990 | 0.000 | 0.845 | 0.018 | NA | NA | NA | novel |
| EIF2B2 | 0.967 | 0.962 | 0.779 | 0.089 | 0.008 | 0.796 | 0.928 | 0.927 | reported |
| KIAA0753 | 0.959 | 0.026 | 0.945 | 0.014 | 0.017 | 0.929 | 0.956 | 0.958 | reported |
| PLEKHJ1 | 0.999 | 0.407 | 0.831 | 0.033 | 0.911 | 0.074 | 0.802 | 0.960 | novel |
| OPRL1 | 0.999 | 0.192 | 0.970 | 0.078 | 0.022 | 0.984 | 0.986 | 0.986 | reported |

These prioritized genes were selected based on the criterion that they showed colocalization with CAD GWAS loci in our primary discovery analysis and passed our specified thresholds in at least two follow-up primary validation analyses. Primary follow-up validation analyses included (a) examination of colocalization with GWAS of subclinical atherosclerosis and (b) investigation of expression artery tissue from GTEx and (c) exanimation of the colocalized genes in PBMC from JHS and whole blood from eQTLGen. Secondary follow-up validation included (a) examination of causal CAD variants for evidence of pQTL and mQTL associations in TOPMed MESA, (b) examination of the association of gene expression with subclinical atherosclerosis traits in MESA and (c) examination of the effects of knockout of the identified gene orthologs in mice **(Figure 1).** We summarize below the results of our primary follow-up validation analyses.

#### Colocalized genes in GWAS of subclinical atherosclerosis

Colocalization analysis leveraging GWAS of subclinical atherosclerosis (CAC^35^ and IMT^36^) and the TOPMed MESA PBMC eQTL resource was performed to investigate the relationship between CAD colocalized genes and subclinical atherosclerosis. Based on this analysis, we identified one CAD colocalized gene, *ZPR1,* showed strong evidence of colocalization with CAC (PP.H4 = 84.5%), and two CAD colocalized gene, *PLEKHJ1* and *ILRUN*, showed strong evidence of colocalization with IMT (*PLEKHJ1*: PP.H4 = 91.1% and *ILRUN*: PP.H4 = 81.5%). **(Figure 3a**, **Table 1 and Table S4)**.

**Figure 3.**
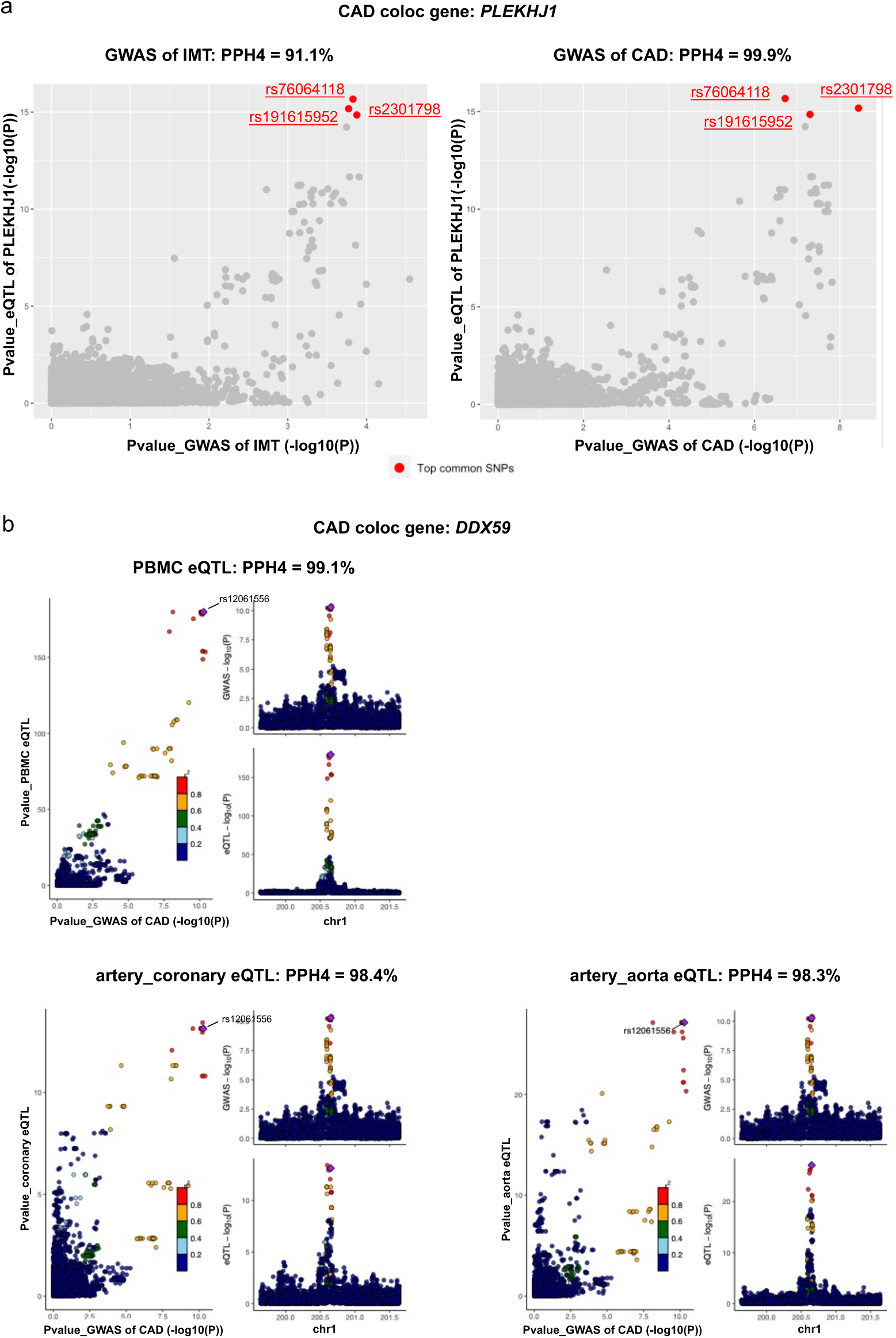
Prioritization of causal genes for CAD and subclinical atherosclerosis from primary follow-up analyses. **Panel a** shows *PLEKHJ1* was identified as a colocalized gene using both GWAS of CAD (right) and IMT (left). **Panel b** shows *DDX59* was identified as a colocalized gene using both eQTL of PBMC (top) and eQTL of coronary (bottom left) and aorta (bottom right).

#### Colocalized genes in artery tissue

CAD-colocalized genes were evaluated for colocalization with eQTLs from GTEx artery tissues (coronary, aorta, tibial). 21 CAD colocalized genes also showed evidence of colocalization in one or more artery tissues **(Table S5)**. For example, *DDX59*, identified based on colocalization with MESA PBMC eQTL, also colocalized with eQTL from GTEx artery tissues (coronary: PP.H4 = 0.984; aorta: PP.H4 = 0.983) **(Figure 3b**, **Table 1 and Table S5).** Additionally, *PLEKHJ1, a* colocalized gene for CAD and IMT, also showed evidence of colocalization in artery tissue (aorta: PP.H4 = 0.802; tibial: PP.H4 = 0.960). These findings indicate the utility of blood-based expression data when vascular tissue is not available.

#### Colocalized genes in PBMC and whole blood tissue

CAD-colocalized genes identified in the primary analysis using MESA PBMC eQTLs were further evaluated for replication using independent eQTL resources, including PBMC eQTLs from JHS and whole-blood eQTLs from eQTLGen. Among the CAD-colocalized genes, 27 showed evidence of replication based on JHS PBMC eQTLs, and 29 showed evidence of replication based on eQTLGen whole-blood eQTLs. For example, *DHDD3*, identified through colocalization with MESA PBMC eQTLs, also demonstrated strong colocalization signals for both JHS PBMCs (PP.H4 = 0.991) and eQTLGen whole blood (PP.H4 = 0.919), supporting the robustness of this association across independent cohorts and expression platforms (**Table 1 and Table S6).**

### Investigation of peak-to-gene links using single-nucleus ATAC-seq (snATAC) dataset

To investigate whether GWAS variants overlapped with regulatory chromatin regions and peak-to-gene links and further evaluate the regulatory relevance of prioritized CAD-colocalized genes, we leveraged the coronary artery snATAC-seq dataset to investigate candidate CAD genes identified through colocalization analyses.

We found that *BICC1* was identified as a marker gene for fibroblasts, *RDX* and *OPRL1* were identified as marker genes for plasma cells, and *CALCRL* was identified as a marker gene for endothelial cells. **(Table S7)** In addition, we examined overlap between CAD-colocalized genes and peak-to-gene (P2G) links, defined as chromatin accessibility peaks whose accessibility correlates with target gene expression. We identified four CAD-colocalized genes (*LIPA*, *DCLRE1B*, *NRIP1*, and *SEMA3C*) that overlapped with P2G links supported by GWAS credible variants (rs11552449, rs1412444, rs2823165, and rs917191), suggesting potential regulatory elements and chromatin interactions that may underlie the observed colocalized eQTL signals. **(Table S7)**

### Identification of causal subclinical atherosclerosis related modules and key driver genes using cWGCNA

cWGCNA was applied to TOPMed MESA PBMC transcriptomic data to investigate network-level regulatory mechanisms underlying subclinical atherosclerosis traits, (CAC, IMT and plaque) in MESA. Analysis of variance indicated that study site and race accounted for the largest proportion of expression variability, followed by sex and age (F statistic > 5), and we included adjustment for these covariates in downstream analyses. Examining associations of the modules with atherosclerosis traits in MESA, module - trait association analysis using limma identified four modules (module8_*STX18-AS1*, module11_*SATB1-AS1*, module16_*TCL6* and module25_*BDH1*) whose eigengenes were statistically significantly associated with plaque, and one module (module28_*PER1*) whose eigengene was statistically significantly associated with IMT after multiple-testing correction (FDR-adjusted p < 0.05 after considering 35 modules). These findings suggest that the expression patterns within the identified modules are related to subclinical atherosclerosis **(Table S8)**.

Within modules significantly associated with subclinical atherosclerosis traits, we further identified key driver genes whose expression levels were independently associated with the corresponding traits in MESA. In summary, we identified 186, 139, 46, 36 and 80 key driver genes in module8_*STX18-AS1*, module11_*SATB1-AS1*, module16_*TCL6*, module25_*BDH1* and module28_*PER1*, respectively. (**Table S9**) For example, 80 genes within the IMT-associated module (module28_*PER1*) showed significant associations with IMT (FDR-adjusted p < 0.05), all exhibiting consistent negative log fold-change values, indicating that lower expression of these genes is associated with increased IMT.

To integrate network-based and colocalization findings, we assessed the overlap between cWGCNA key driver genes within subclinical atherosclerosis–associated modules and CAD-colocalized genes identified from MESA PBMC eQTL analyses. Three genes within plaque-associated modules - *ATG9B* in module11_*SATB1-AS1* and *PRAM1* and *ZBTB46* in module8_*STX18-AS1* - were also among the CAD-colocalized genes, demonstrating the utility of combining colocalization with co-expression network analysis to prioritize key driver genes **(Table 2).** We further compared the PBMC cWGCNA key driver genes with CAD-colocalized genes identified from CD4+ T-cell and monocyte colocalization analyses. Two PBMC key driver genes (*PRAM1* and *ZBTB46*) were also identified as CAD-colocalized genes in CD4+ T cells, while one PBMC key driver gene (*ABI3*) was also identified as a CAD-colocalized gene in monocytes. These findings suggest that several prioritized genes are supported by both network-based and regulatory genetic evidence across multiple immune cell populations, highlighting their potential relevance to CAD and subclinical atherosclerosis.

**Table 2.** Overlap investigation between CAD-colocalized genes identified from MESA PBMC eQTL analyses and key driver genes within subclinical atherosclerosis–associated modules identified from cWGCNA.

| Module | Key driver gene | logFC | p.value | adj.p.value | PPH4_CAD |
| --- | --- | --- | --- | --- | --- |
| module11_SATB1-AS1 | ATG9B | -0.135 | 0.021 | 0.038 | 0.821 |
| module8_STX18-AS1 | PRAM1 | 0.146 | 0.012 | 0.039 | 0.842 |
| module8_STX18-AS1 | ZBTB46 | 0.117 | 0.021 | 0.049 | 0.959 |

### Pathway enrichment analysis for identification of biological pathways for subclinical atherosclerosis related modules

Gene Set Enrichment Analysis was performed to investigate the relevant pathways for subclinical atherosclerosis related modules (module8_*STX18-AS1*, module11_*SATB1-AS1*, module16_*TCL6*, module25_*BDH1* and module28_*PER1*) identified by cWGCNA, leveraging the Molecular Signatures Database (MSigDB). One of IMT-related module, module28_*PER1*, identified significant downregulation of hypoxia and TNF-α/NF-κB signaling pathways (FDR-adjusted p < 0.05). These findings suggest Module28 reflects a distinct transcriptional profile associated with inflammatory and hypoxic signaling **(Table S10)**

## Discussion

While previous GWAS of CAD have made significant progress by identifying over 300 independent genetic loci associated with CAD, systematic prioritization of candidate genes and comprehensive investigation of their relationships with subclinical atherosclerosis remain limited. To address this critical gap, the present study aimed to elucidate the genetic mechanisms linking CAD with subclinical atherosclerosis by integrating multi-omics data from the TOPMed MESA cohort with multi-ancestry GWAS data for CAD and subclinical atherosclerosis traits. We applied Bayesian colocalization analysis - with and without statistical fine-mapping - followed by a series of follow-up validation analyses to prioritize 5 novel (*CCDC30, ZEB1-AS1, ZPR1, PLEKHJ1 and AC018816.3*) and 8 previously reported genes (*DHDDS, DDX59, LNPEP, DAGLA, ZKSCAN1, LIPA, OPRL1 and EIF2B2*) with putative roles in both CAD and subclinical atherosclerosis. Additionally, we utilized cWGCNA to identify 5 modules of genes exhibiting significant association with subclinical atherosclerosis (IMT and Plaque), allowing us to explore potential pathways linked to these genes in greater depth. We further identified that three genes within plaque-associated modules - *ATG9B* in Module11 and *PRAM1* and *ZBTB46* in Module8 - were also identified as CAD-colocalized genes.

Among the candidate genes prioritized from colocalization analysis and follow-up validation analysis, recent functional evidence suggests that reduced *ZEB1* in macrophages increases atherosclerotic plaque formation in Apoe-knockout mice, with more lipid accumulation in macrophages due to delayed lipid traffic and deficient cholesterol efflux. In addition, in human endarterectomy samples, lower *ZEB1* expression is associated with plaque rupture and cardiovascular events, supporting relevance to human disease.^47^ Given that *ZEB1-AS1,* the novel CAD prioritized gene, is an antisense transcript at the *ZEB1* locus, our colocalization evidence implicating *ZEB1-AS1* is consistent with the possibility that regulatory variation of the *ZEB1* contributes to CAD and subclinical atherosclerosis through immune-cell mechanisms. Prior studies also suggest that *ZEB1-AS1* can modulate responses to atherosclerosis-relevant stimuli (e.g., ox-LDL–related endothelial injury), providing additional biological plausibility for this locus.^48^

Mendelian randomization (MR) is a widely used framework for causal inference but its application to gene expression traits can be challenging in practice. MR relies on the availability of valid instrumental variables that are strongly associated with the exposure and influence the outcome exclusively through that exposure. For gene expression, these assumptions are often difficult to satisfy because regulatory variants frequently influence multiple genes or biological pathways and may affect disease risk through mechanisms that are not mediated by the exposure of interest, which can violate key assumptions of Mendelian randomization.^49,50^ In contrast, Bayesian colocalization analysis, one of integrative analysis, is specifically designed to evaluate evidence of shared genetic variant(s) underlying both a molecular trait and a disease association within a locus, without requiring explicit instrumental variable selection.

Our study incorporated both Bayesian colocalization analysis with and without statistical fine-mapping to identify a total of 108 genes showing evidence of colocalization with CAD loci. Our results illustrate the value of incorporating statistical fine-mapping into the Bayesian colocalization analysis.^29^ While some genes were readily identified in colocalization analyses without fine-mapping, the complementary colocalization approach incorporating fine-mapping allowed us to address the scenario of multiple independent signals in a specific GWAS region (typically defined as the transcript start site +/− 1Mb). For example, *SCARB1,* one of reported colocalized gene of CAD, was identified by incorporating statistical fine-mapping into colocalization analysis. *S*tatistical fine-mapping identified multiple credible sets for both the GWAS of CAD and the eQTL signals. On the other hand, it is worth noting that in situations where the sample size or the statistical power of GWAS and eQTL does not allow for the confident detection of credible sets by SuSiE, basic colocalization analysis under the single causal variant assumption can still play a valuable role in identifying colocalized genes.^26^ Three follow-up validation analyses in our study, colocalization analysis using GWAS of subclinical atherosclerosis (CAC and IMT), colocalization analysis using eQTL from GTEx artery tissues and colocalization analysis using JHS PBMC eQTL and eQTLgen whole blood eQTL were performed by basic colocalization analysis under the single variant assumption, as the relatively small sample size limited our ability to apply statistical fine-mapping.

Another distinguishing feature of our study is the use of multi-ancestry GWAS of CAD and multi-ancestry MESA eQTL resources, with both resources having been constructed using data sets spanning individuals of European, African, Hispanic and Asian race/ancestry. To date, most statistical genetic studies have focused primarily on a single ancestry group, predominantly emphasizing European ancestry. This single ancestry approach can be underpowered to detect certain genetic signals due to differences in allele frequencies, LD patterns and effect sizes across ancestries.^51–54^ The incorporation of multi-ancestry resources in our study conferred several noteworthy advantages. For example, use of multi-ancestry resources allowed for increased sample sizes and broader representation of multi-ancestry LD structures by incorporating participants from different ancestry groups, which further enhanced the statistical power and resolution of our subsequent fine-mapping and colocalization analyses.

In this study, we performed colocalization analyses using MESA PBMC-derived eQTLs and further evaluated prioritized genes using independent eQTL resources from JHS PBMCs, eQTLGen whole blood, and GTEx artery tissues. A subset of CAD-colocalized genes identified in the primary MESA PBMC analysis showed consistent colocalization signals across these independent datasets, supporting the reproducibility and biological relevance of these associations. For example, *DDX59* showed evidence of colocalization across GTEx artery tissues, JHS PBMC eQTLs, and eQTLGen whole-blood eQTLs, supporting its potential role in CAD-related regulatory mechanisms. Our findings, therefore, support the feasibility of using blood-derived molecular data to uncover genetic mechanisms underlying CAD and provide a rationale for leveraging readily available blood-based omics in large-scale cardiovascular genomics research research.^55,56^

At the same time, we observed differences between PBMC- and whole-blood– based colocalization results, likely reflect underlying biological and cellular composition differences. PBMC transcriptomes represent a defined immune-cell subset of peripheral blood and are enriched for immune and inflammatory regulatory mechanisms, whereas whole-blood transcriptomes capture a heterogeneous mixture of cell types and are strongly influenced by cell-type composition effects.^57^ In addition, differences in population ancestry between MESA and JHS may also contribute to variability in colocalization results. The MESA eQTL resource used in the primary analysis is derived from a multi-ancestry cohort, whereas the JHS eQTL dataset consists primarily of African American participants. Differences in allele frequencies, linkage disequilibrium structure, and genetic architecture across populations can influence both eQTL detection and colocalization signals, potentially leading to population-specific regulatory associations and incomplete concordance between datasets.^58,59^ Together, these findings highlight both the utility and limitations of blood-derived eQTL resources and emphasize the importance of considering tissue and cellular context when interpreting colocalization results.

Although colocalization analysis and follow-up validation analysis prioritize multiple candidate genes with putative roles in both CAD and subclinical atherosclerosis, not all disease-relevant genes are expected to be genetically regulated at the transcriptional level. ^60–63^ Gene expression and protein activity can also be influenced by post-transcriptional and post-translational regulatory mechanisms, including regulation by microRNAs, RNA-binding proteins, alternative splicing, and mRNA stability, as well as protein-level modifications such as phosphorylation, acetylation, methylation, glycosylation, and ubiquitination. ^60–63^ These mechanisms may contribute to disease processes but may not be captured by eQTL-based analyses. To further evaluate the regulatory relevance of prioritized CAD-colocalized genes, we incorporated coronary artery snATAC-seq data to examine whether GWAS credible variants overlapped regulatory chromatin regions and peak-to-gene (P2G) links, four CAD-colocalized genes (*LIPA, DCLRE1B, NRIP1, and SEMA3C*) overlapped with P2G links supported by GWAS credible variants, suggesting that these variants may influence CAD risk through regulatory chromatin interactions affecting target gene expression.

Additionally, our study applied causal weighted gene co-expression network analysis (cWGCNA) leveraging the TOPMed MESA PBMC transcriptomics data to identify gene co-expression modules and key driver genes associated with subclinical atherosclerosis traits. We compared cWGCNA-derived key driver genes with CAD-colocalized genes identified from PBMC eQTL analyses. Notably, three key driver genes (*ATG9B*, *PRAM1* and *ZBTB46*) identified by cWGCNA were also identified as CAD-colocalized genes. *ATG9B* (Autophagy Related 9B) encodes an autophagy-related protein involved in vesicle trafficking, and dysregulation of autophagy has been linked to endothelial dysfunction, atherosclerotic plaque development, and cardiovascular homeostasis. In a case–control study of angiographically confirmed CAD patients, polymorphism rs2373929 in *ATG9B* showed a strong association with CAD risk, with individuals carrying the TT genotype exhibiting a significantly increased risk compared with controls (odds ratio ≈3.6, P < 0.001).^64^ Together with our findings, these results suggest *ATG9B* may represent a biologically plausible candidate gene involved in CAD risk. *ZBTB46* (Zinc Finger and BTB Domain Containing 46) is a repressive transcription factor and a widely accepted marker for classical dendritic cells (DCs). Experimental studies show *ZBTB46* is expressed in endothelial cells and classical dendritic cells, influences endothelial proliferation under shear stress, and thus may contribute to vascular homeostasis and response to disturbed flow - a key atherogenic stimulus.^65^

Despite its strengths, our study also faces several limitations. Colocalization analyses were primarily conducted using PBMC-derived eQTL data in this study, which capture gene regulatory effects in circulating immune cells. Many of the genes most strongly associated with CAD in large GWAS are known to act through mechanisms related to lipid metabolism, vascular smooth muscle function, or arterial wall biology, which may not be well represented in PBMC transcriptomes. As a result, such genes are not necessarily expected to exhibit regulatory effects in blood-derived expression datasets. To address this limitation, we applied the follow-up validation analysis using eQTL from GTEx Artery tissue. 21 CAD-colocalized genes identified using PBMC eQTLs also showed evidence of colocalization with eQTLs from GTEx artery tissues. However, to further enhance the relevance and comprehensiveness of our discovery analysis, it would be advantageous to leverage eQTL data from other disease-related tissues available in existing databases. For instance, the Stockholm-Tartu Atherosclerosis Reverse Network Engineering Task (STARNET) study represents a valuable eQTL resource for our primary analysis.^66^ Additionally, although colocalization provides evidence that genetic variation at CAD colocalized locus influences both gene expression and CAD risk, direct evaluation of gene expression of CAD colocalized genes in disease-relevant arterial tissues with clinical outcome was beyond the scope of this study and remains an important area for future investigation. Additionally, multiple recent studies have demonstrated that positive predictive value of eQTLs in identifying causal genes for complex diseases is limited. Most disease-associated variants, despite being located in putatively regulatory regions, do not show detectable effects on gene expression. Moreover, the regulatory variants identified from eQTLs account for only a small fraction of the signals detected in GWAS.^67–69^ To address this limitation, we performed a correlation network analysis, cWGCNA, to identify subclinical atherosclerosis associated modules by using the TOPMed MESA transcriptomics data and utilized Molecular Signature Database (MSigDB) to explore the pathways among genes within the modules of interest.

An additional limitation of this study is that we did not evaluate the predictive performance of network-derived modules or prioritized gene sets in independent cohorts. Such analyses require external datasets that include matched transcriptomic dataset and detailed subclinical atherosclerosis phenotypes (e.g., CAC or IMT), which are currently limited. Future studies leveraging larger cohorts may enable assessment of the predictive utility of these gene modules and further clarify their translational relevance. Furthermore, future studies integrating fine-mapped variants with variant-based prediction methods may provide additional insight into regulatory mechanisms linking genetic variation to CAD risk.^70–72^ Finally, although our integrative genomic analyses prioritize candidate genes, another limitation of this study is that we did not have access to arterial tissue expression data from the MESA participants. As a result, we were unable to directly assess whether expression of prioritized genes. Future studies using artery-specific transcriptomic data and experimental validation will be needed to further characterize the functional role of these genes in CAD.

In summary, our study employed a diverse array of statistical analyses leveraging the transcriptomics data from TOPMed MESA to prioritize candidate genes associated with CAD and subclinical atherosclerosis. In the future, we anticipate that follow-up experimental validation focusing on the candidate genes of CAD we identified can be conducted to advance understanding of these prioritized candidates. Collectively, these findings underscore the value of incorporating statistical fine-mapping in colocalization studies and demonstrate the utility of combining colocalization with co-expression network analysis to prioritize functional genes, which provide a better understanding of genetic mechanisms and pathways implicated by GWAS of CAD and subclinical atherosclerosis and consequently provide valuable insights into potential therapeutic interventions and treatments.

## Data Availability

Individual whole-genome sequence data for TOPMed whole genomes are available through dbGaP. The dbGaP accession numbers of Multi-Ethnic Study of Atherosclerosis (MESA) is phs001416. Data in dbGaP can be downloaded by controlled access with an approved application submitted through their website: https://www.ncbi.nlm.nih.gov/gap.

## Acknowledgments

This work was supported by a Predoctoral Fellowship from the American Heart Association (C.Y.). We acknowledge funding support from the National Institutes of Health (NIH) / National Heart Lung and Blood Institute (NHLBI) grants: R01HL148239 and R01HL164577 (C.L.M.). The authors acknowledge Research Computing at The University of Virginia for providing computational resources and technical support that have contributed to the results reported within this publication.

## The Multi-Ethnic Study of Atherosclerosis

The Multi-Ethnic Study of Atherosclerosis (MESA) projects are conducted and supported by the National Heart, Lung, and Blood Institute (NHLBI) in collaboration with MESA investigators. Support for MESA is provided by contracts 75N92020D00001, HHSN268201500003I, N01-HC-95159, 75N92020D00005, N01-HC-95160, 75N92020D00002, N01-HC-95161, 75N92020D00003, N01-HC-95162, 75N92020D00006, N01-HC-95163, 75N92020D00004, N01-HC-95164, 75N92020D00007, N01-HC-95165, N01-HC-95166, N01-HC-95167, N01-HC-95168, N01-HC-95169, UL1-TR-000040, UL1-TR-001079, UL1-TR-001420, UL1TR001881, DK063491, and R01HL105756. The authors thank the other investigators, the staff, and the participants of the MESA study for their valuable contributions. A full list of participating MESA investigators and institutes can be found at http://www.mesa-nhlbi.org.

Whole Genome Sequencing (WGS) for “NHLBI TOPMed: Multi-Ethnic Study of Atherosclerosis (MESA)” (phs001416) was performed at the Broad Institute of MIT and Harvard (3U54HG003067-13S1 and HHSN268201500014C). RNA-seq for the NHLBI TOPMed: Multi-Ethnic Study of Atherosclerosis (MESA)” (phs001416.v1.p1) was performed at the Northwest Genomics Center (HHSN268201600032I) and at the Broad Institute Genomics Platform (HHSN268201600034I). Genome-wide methylation for NHLBI TOPMed: Multi-Ethnic Study of Atherosclerosis (MESA)” (phs001416.v1.p1) was performed at Keck Molecular Genomics Core Facility (HHSN268201600034I). SOMAscan proteomics for NHLBI TOPMed: Multi-Ethnic Study of Atherosclerosis (MESA)” (phs001416.v1.p1) was performed at the Broad Institute and Beth Israel Proteomics Platform (HHSN268201600034I). Centralized read mapping and genotype calling, along with variant quality metrics and filtering were provided by the TOPMed Informatics Research Center (3R01HL-117626-02S1; contract HHSN268201800002I). Phenotype harmonization, data management, sample-identity QC, and general study coordination, were provided by the TOPMed Data Coordinating Center (3R01HL-120393-02S1, contract HHSN268201800001I), and TOPMed MESA Multi-Omics (HHSN2682015000031/HSN26800004). We gratefully acknowledge the studies and participants who provided biological samples and data for TOPMed. The views expressed in this manuscript are those of the authors and do not necessarily represent the views of the National Heart, Lung, and Blood Institute; the National Institutes of Health; or the U.S. Department of Health and Human Services. A full list of investigators for the NHLBI Trans-Omics for Precision Medicine (TOPMed) Consortium is provided at https://topmed.nhlbi.nih.gov/topmed-banner-authorship.

## RNA-Seq eQTL

Freeze 1RNA TOPMed cis-eQTL results were generated in a collaboration between the TOPMed Informatics Research Center, TOPMed Multi-Omics working group, and the TOPMed parent studies contributing RNA-seq and distributed to TOPMed investigators. We acknowledge the contributing cohorts, sequencing centers, and the TOPMed IRC.

