## Supplementary text for "Systematic Integration of genomics with transcriptomics for the Study of Coronary Artery Disease and Subclinical Atherosclerosis"

**Correspondence:**

Ani Manichaikul, PhD

Associate Professor of Genome Sciences

The total word count of the supplementary text: 2031

### Supplementary methods

### Secondary follow-up analyses

#### Association of colocalized genes with subclinical atherosclerosis

CAD-colocalized genes identified from primary analysis were carried forward to examine the association with subclinical atherosclerosis traits (CAC, IMT and carotid plaque) in MESA exam 5 using linear regression model and the covariates included age, sex and study sites. A nominal p-value (0.05) was used as the threshold of association. In MESA exam5, CAC was measured with either electron-beam computed tomography (EBT) at 3 field centers or multidetector computed tomography (MDCT) at 3 field centers.[1–3] The amount of calcium was quantified with the Agatston scoring method. Carotid IMT was defined as the intima-media thickness measured as the mean of the left and right far wall distal common carotid artery wall thicknesses. Carotid plaque was defined as a discrete, focal wall thickening ≥1.5 cm or focal thickening at least 50% greater than the surrounding IMT.

#### Examination of causal CAD variants for evidence of pQTL and mQTL associations for the corresponding colocalized genes

Shared causal variants were identified by statistical fine-mapping (SuSIE) in GWAS of CAD and MESA eQTL from the primary analysis. These shared causal variants were further investigated to assess their potential impact as MESA pQTL or MESA mQTL for the proteins and CpG sites corresponding to CAD-colocalized genes. TOPMed MESA multi-ancestry pQTL resources included 1,305 proteins measured by a SOMAscan assay and 971 unique individuals (African American [n = 183], Chinese [n = 71], European [n = 416], and Hispanic/Latino [n = 301]).[8] TOPMed MESA multi-ancestry mQTL resources included the whole blood DNA methylation (DNAm) data for 747,868 CpG sites (CpG sites passing QC - 740,291) and 900 unique individuals.[9]

**Investigation of CAD colocalized genes in mouse genome**

To investigate the biological function of CAD-colocalized genes in mouse genome, we utilized the data from the International Mouse Phenotyping Consortium (IMPC)[10] and Mouse Genome Informatics (MGI)[11] to determine whether these CAD-colocalized genes are associated with heart/cardiovascular phenotypes, for example, heart morphology, heart rate and cardiovascular system phenotypes. IMPC is an international effort by 21 research institutions, consisting of 85M data points and over 95,000 statistically significant phenotype hits mapped to human disease, to identify the function of every protein-coding gene in the mouse genome. MGI is the international database resource for the laboratory mouse, providing integrated genetic, genomic, and biological data to facilitate the study of human health and disease.

### Supplementary results

### Secondary follow-up analyses

Secondary follow-up included (a) examination of causal CAD variants for evidence of pQTL and mQTL associations for the corresponding colocalized genes in TOPMed MESA, (b) examination of the association of colocalized genes with subclinical atherosclerosis traits in MESA and (c) study of the colocalized genes in mouse genome.

**Association of colocalized genes with subclinical atherosclerosis:** Linear regression analysis was performed to examine the association between measured expression of CAD colocalized genes and subclinical atherosclerosis traits (CAC, IMT, and carotid plaque) in MESA. Nominal significance was observed for multiple genes (e.g., *CAMSAP2, MED19, CNPY2*, *ALDH2, LAYN* and *GGCX*; **Table S11)**.

**pQTL and mQTL associations of colocalized genes:** Overlapping causal variants, identified by statistical fine-mapping in GWAS of CAD and eQTL, were examined for evidence supporting them as pQTLs (proteins) or mQTLs (CpG sites of DNA methylation) corresponding to CAD-colocalized genes. rs11213945, one of the causal variants associated with CAD and gene expression of *LAYN*, had strong significant association with the corresponding protein (LAYN) as a pQTL and with the CpG site, cg21703322, as a mQTL **(Figure S2)**.

**Colocalized genes in mouse genome:** Three CAD colocalized genes (*SCARB1*, *BICC1*, and *PAN2*) showed significant associations with heart and cardiovascular-related functions in the mouse genome, using data from the International Mouse Phenotyping Consortium (IMPC) and Mouse Genome Informatics (MGI). Mouse knockouts for *SCARB1* had decreased heart rate, abnormal sinus arrhythmia, and cardiovascular system traits (e.g., observable morphological and physiological characteristics of the mammalian heart, blood vessels, or circulatory system that are manifested through development and lifespan). **(Table S12)**

### Competing Interests

The authors declare no competing interests.

**Supplementary Figure Legends**

**Figure S1** shows the identification of colocalized genes for CAD across different cell types. **Panel a** shows the colocalized genes for CAD identified using eQTL from TOPMed MESA for PBMCs, T cells and monocytes. Panel b shows the sample size of eQTL for PBMCs, T cells and monocytes.

**Figure S2** shows the shared causal variant has strong associations in GWAS of CAD, eQTL, pQTL and mQTL for *LAYN*.

### Figure S1. Identification of colocalized genes for CAD across different cell types.

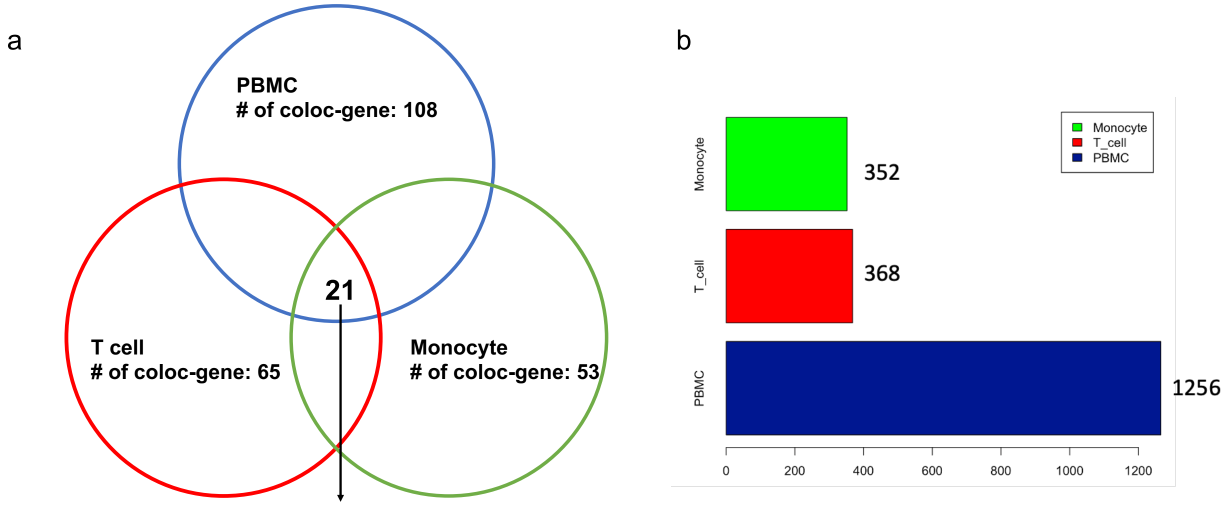


CAD-colocalized gene identified across all cell types:*FHL3, UTP11, CCDC30, DDX59, RP11-378J18.8, PRKD3, USP34-DT, GFPT1, ILRUN, NHSL1, DAGLB, BICC1, LIPA, MED19, CWF19L2, EIF2B2, TOM1L2, MRPL45P2, VN1R84P, ZNF100, PHF23*

**Figure S1** shows the identification of colocalized genes for CAD across different cell types. **Panel a** shows the colocalized genes for CAD identified using eQTL from TOPMed MESA for PBMCs, T cells and monocytes. Panel b shows the sample size of eQTL for PBMCs, T cells and monocytes.

### Figure S2. Examination of causal CAD variants for evidence of pQTL and mQTL associations for the corresponding colocalized genes

**CAD coloc gene: *LAYN* (PPH4 = 0.955)**

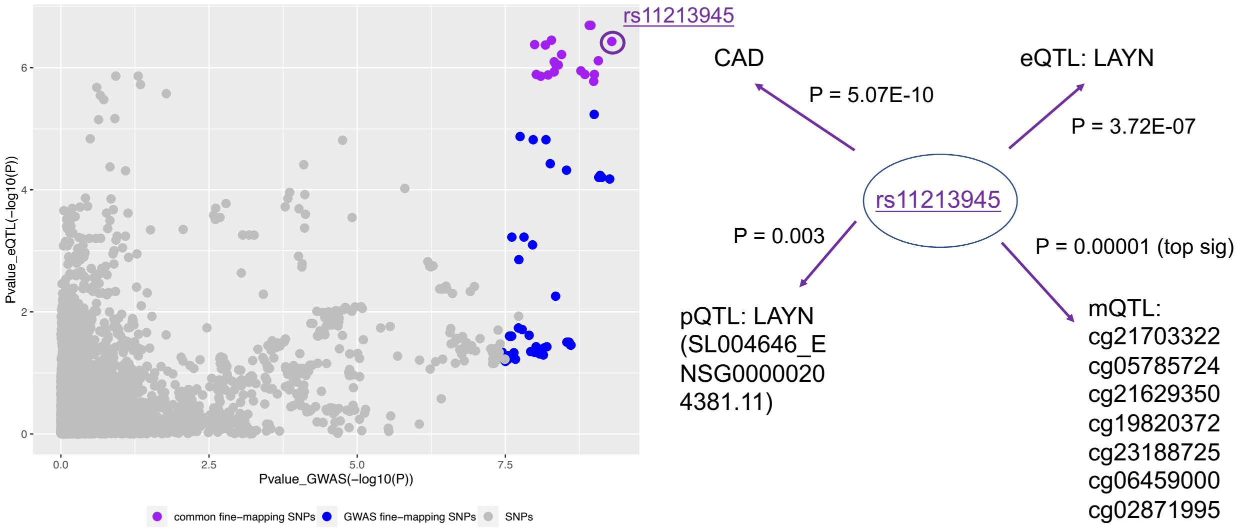


Figure S2 shows the shared causal variant has strong associations in GWAS of CAD, eQTL, pQTL and mQTL for *LAYN*.
